# Management of Penetrating Injuries of Abdomen and Its Outcome

**DOI:** 10.1101/2025.09.27.25336795

**Authors:** Deepika Shelar, Anitha Kandi, Diksha Srivastava, Sarojini Jadha

## Abstract

**Objective:** To evaluate the clinical profile, management, and outcomes of penetrating abdominal injury.

**Background:** Penetrating abdominal trauma is a leading cause of morbidity and mortality worldwide. Prompt recognition and timely intervention are crucial to patient survival.

**Methods:** This prospective observational study was conducted at a tertiary care government center between September 2020 and October 2022. Patients with penetrating abdominal injuries were evaluated by detailed history, clinical examination, and laboratory and radiological investigations and managed according to standard protocols. The operative findings and postoperative outcomes were also recorded.

**Results:** A total of 68 patients with penetrating abdominal trauma were studied, with a marked male predominance and peak incidence in the 21–30 year age group. Homicidal stab wounds were the leading cause (85%), while other causes included bullhorn injuries, self-inflicted wounds, and accidental trauma (5% each). Clinical presentations were dominated by peritoneal penetration (67%), omental evisceration (32%), and bowel evisceration (10%). Radiological evaluation revealed diagnostic utility of CECT (50% positivity) compared to FAST (29%) and plain radiography (22%). The mesentery (36%), small bowel (34%), and colon (25%) were the most commonly injured organs. Primary repairs of mesentery (42.5%), small bowel (37.5%), and colon (17.5%) were the main surgical interventions, with diversion stomas required in selected cases. In 13% of patients, no visceral injury was found. Mean hospital stay was 7 days, with an overall mortality of 13.2%. Postoperative complications included respiratory infections, ARDS, abdominal sepsis, surgical site infections, and late complications such as burst abdomen, stoma prolapse, and anastomotic leakage.

**Conclusions:** Penetrating abdominal injuries predominantly affect young adults and are associated with significant morbidity and mortality rates. Early diagnosis, appropriate resuscitation, and timely surgical intervention are cornerstones of management.

## Introduction

Trauma is still the most frequent cause of death in the first four decades of life, remains a major public health problem in every country, regardless of the level of socioeconomic development, and is the third most common cause of death regardless of age. It is more common in males than in females. Abdomen is the diagnostic black box. It is the third most commonly injured region, with surgery required in approximately 25% of civilian cases.^1^

The mortality rate was very high in patients with penetrating abdominal injuries. Among the various modes of trauma, penetrating trauma requires immediate surgical intervention in the majority of cases.^2^ As most of the deaths due to penetrating injuries occur within minutes to hours, they form an important part of surgical emergencies.

The most common organs involved in penetrating abdominal trauma are the small intestine and colon, and most postoperative complications are related to these organs.^3^

Multiorgan injuries, exsanguinating hemorrhage, delayed presentations, and the ominous reputation for high mortality and morbidity are just a few of the many reasons that make this topic of penetrating injuries fascinating. Appropriate and expeditious assessment and investigation of individuals with penetrating abdominal injuries facilitates definitive management and minimizes the risk of complications.□

Adjunctive diagnostic testing, including ultrasonography, computed tomography, local wound exploration, diagnostic peritoneal lavage, and laparoscopy, is often used to identify significant injuries requiring operative management.□There are a number of factors that determine outcome in penetrating abdominal trauma; therefore, the aim of this study was to study the management of penetrating injuries of abdomen and its outcome in a tertiary care center.

## Materials and Methods

This prospective observational study was conducted at a tertiary care center in a government set up over a period of two years from September 2020 to October 2022, after approval by the Institutional Ethical Committee. Patients who presented with trauma were screened, and those with penetrating injuries to the abdomen as per the inclusion criteria were enrolled in the study after obtaining written informed consent.

A detailed clinical history was obtained from all patients, followed by a thorough clinical examination. They were further investigated by haematological and radiological investigations. A detailed primary and secondary survey was conducted, and accordingly, patients underwent the operative procedure after resuscitation. The patients were monitored closely for postoperative complications.

### Inclusion criteria

All patients with penetrating injury to abdomen.

### Exclusion criteria

- All patients with penetrating abdominal injuries who were dead on arrival.
- All patients with blunt trauma to abdomen.
- All patients had penetrating trauma other than the abdomen (i.e., exclusively to the chest, extremities, head, neck, and face).
- Patients not willing to take part in study.

## Results

A total of 68 patients with penetrating abdominal trauma were included in the study. The cohort demonstrated a marked male predominance, with the highest incidence observed among individuals aged 21–30 years, reflecting the vulnerability of young adults to interpersonal violence and occupational hazards.

### Mode of injury

Homicidal stab wounds represented the leading cause of injury (56 patients, 85%), while bullhorn injuries, self-inflicted wounds, and accidental trauma each accounted for 5% (n=4) of cases.

### Clinical presentation

The predominant presentation was peritoneal penetration (46 patients, 67%), followed by omental evisceration (22 patients, 32%) and bowel evisceration (7 patients, 10%). A subset presented with combined omental and bowel evisceration (5 patients, 7%) or hemodynamic instability (5 patients, 7%).

### Diagnostic evaluation

Radiological investigations varied across patients. An erect abdominal radiograph was performed in 36 patients, yielding significant findings in 8 cases (22%). Focused Assessment with Sonography for Trauma (FAST) was carried out in 31 patients, identifying free fluid or organ injury in 9 cases (29%). Contrast-enhanced computed tomography (CECT) was selectively performed in 10 patients, with positive findings in 5 (50%), underlining its value in hemodynamically stable patients.

### Injury profile and operative management

The mesentery (36%), small bowel (34%), and colon (25%) were the most frequently injured structures. Less common injuries involved the omentum (20%), vascular structures (10%), stomach (7%), liver (2%), spleen (2%), gallbladder (2%), and urinary bladder (2%).

The predominant procedures included mesenteric repair (42.5%), primary small bowel repair (37.5%), and primary colonic repair (17.5%). Diversion stomas (ileostomy/colostomy/sigmoidostomy) were performed in selected cases (total 10%).

Omental repairs accounted for 17% of procedures, while vascular, gastric, hepatic, splenic, gallbladder, and bladder repairs constituted the remainder. In 13% of patients (n=6), no visceral injury was identified, and peritoneal lavage with drainage was sufficient.

### Hospital course and outcomes

The mean hospital stay was 7 days (range 4–18 days). The overall mortality rate was 13.2% (n=9), predominantly due to exsanguination, septic complications, and multi-organ dysfunction.

### Complications

Early postoperative morbidity included respiratory infections and acute respiratory distress syndrome (ARDS), abdominal sepsis, and surgical site infections. Late complications comprised burst abdomen, stoma prolapse, and anastomotic leakage.

## Discussion

Several studies have reported findings similar to those of this study. Patel et al. observed that young adults were the most affected by penetrating abdominal trauma. Ramya et al. and Tillu et al.^3^ also reported similar age distributions and sex predominance.

Rachha et al.^8^ and Omer et al.^1^ emphasized the mesentery and small intestine are the most frequently injured organs, which matches the findings of this study as per table 4.

**Table 1:** Mode of penetrating injury.

| Mode of injury | Number of patients | Percentage(n=68) |
| --- | --- | --- |
| Homicidal stab injury | 56 | 85 |
| Bull horn injury | 4 | 5 |
| Self inflicted stab injury | 4 | 5 |
| Accidental injuries | 4 | 5 |
| Total | 68 | 100 |

**Table 2:** Clinical Presentation.

| Clinical presentation | Number of patients | Percentage(n=68) |
| --- | --- | --- |
| Peritoneal penetration | 46 | 67 |
| Omental evisceration | 22 | 32 |
| Bowel Evisceration | 7 | 10 |
| Omental with bowel evisceration | 5 | 7 |
| Hemodynamic instability | 5 | 7 |

**Table 3:** Radiological Investigations.

| Investigations | Done | Not done | Significant findings |
| --- | --- | --- | --- |
| Xray abdomen erect (n=68) | 36 | 32 | 8 |
| Ultrasonography (FAST) (n=68) | 31 | 37 | 9 |
| Computed tomography(CT) (n=68) | 10 | 58 | 5 |

**Table 4:** Injured organ and operative procedure.

| Organ | Number of patients | Percentage (n=46 ) | Operative procedure | Number of patients | Percentage (n=46) |
| --- | --- | --- | --- | --- | --- |
| Small bowel | 16 | 34 | -Primary repair | 15 | 37.5 |
|  |  |  | -Ileostomy | 1 | 2.5 |
| Mesentery | 17 | 36 | Mesenteric repair | 17 | 42.5 |
| Colon | 10 | 25 | -Primary repair | 7 | 17.5 |
|  |  |  | -Transverse colostomy | 1 | 2.27 |
|  |  |  | -Sigmoidostomy | 2 | 5 |
| Omentum | 8 | 20 | Omental repair | 8 | 17 |
| Vascular | 5 | 10 | Vascular repair and ligation | 5 | 10 |
| Stomach | 3 | 7 | Gastric perforation repair | 3 | 7.5 |
| Liver | 1 | 2 | Hemostat application in liver laceration | 1 | 2.5 |
| Spleen | 1 | 2 | Splenorrhaphy | 1 | 2.5 |
| Gall bladder | 1 | 2 | Cholecystectomy | 1 | 2.5 |
| Urinary bladder | 1 | 2 | Urinary bladder repair | 1 | 2.5 |
| No visceral organ injury | 6 | 13 | Peritoneal lavage and drainage | 6 | 13 |

The complication rates in our study were consistent with those reported in the literature, including respiratory complications, surgical site infections, and abdominal sepsis.^9,10^

## Conclusion

Penetrating abdominal trauma is a leading cause of morbidity and mortality. The most common penetrating injuries in civilian groups involve younger, healthy populations, who are responsible for the development of society. The establishment of a coordinated trauma registry and system of care and evacuation teams is highly recommended in our setting and in developing countries to curb the prevailing morbidity and mortality rates. Hemodynamically stable patients and those without signs of peritonitis should be evaluated further; thus, facilities for diagnostic laparoscopy and selective angiography should be made available during emergency hours. This would result in a shorter hospital stay and reduce mortality and morbidity associated with unnecessary surgical intervention. Additional multicenter studies with a greater number of patients for a longer period are needed to substantiate the preliminary evidence of this study.

**Fig 1a:**
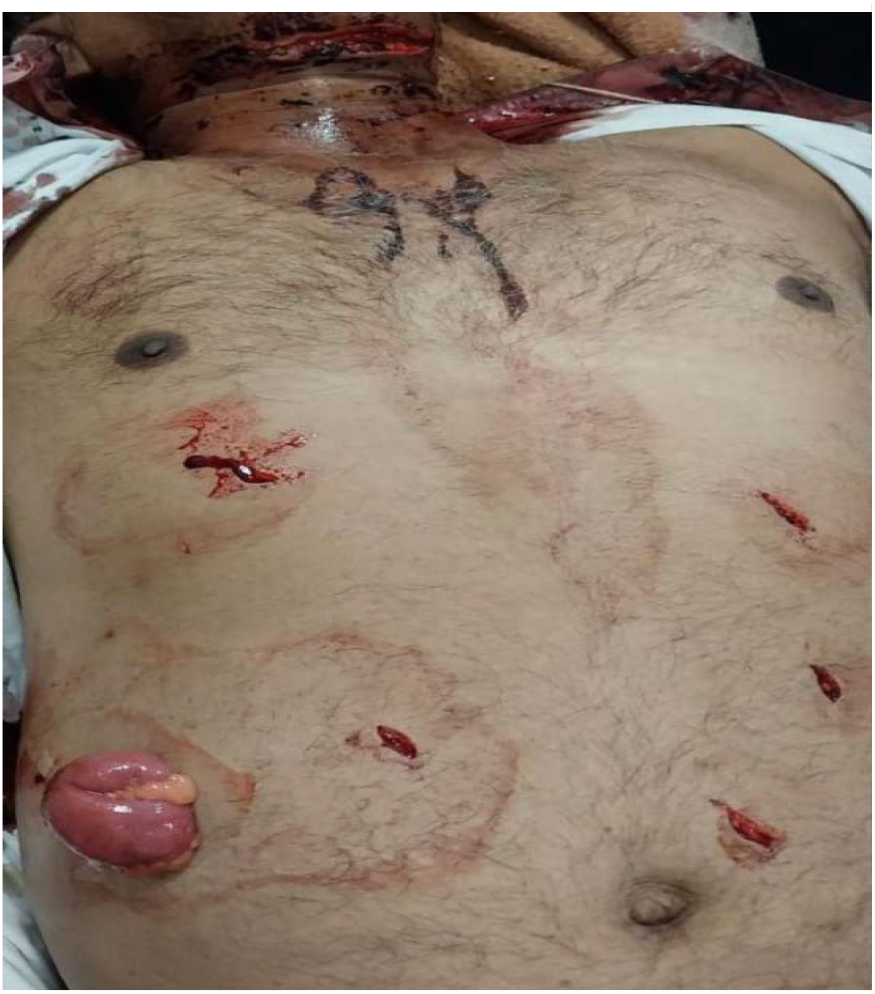
Multiple stab wounds over abdomen with evisceration of bowel.

**Fig 1b:**
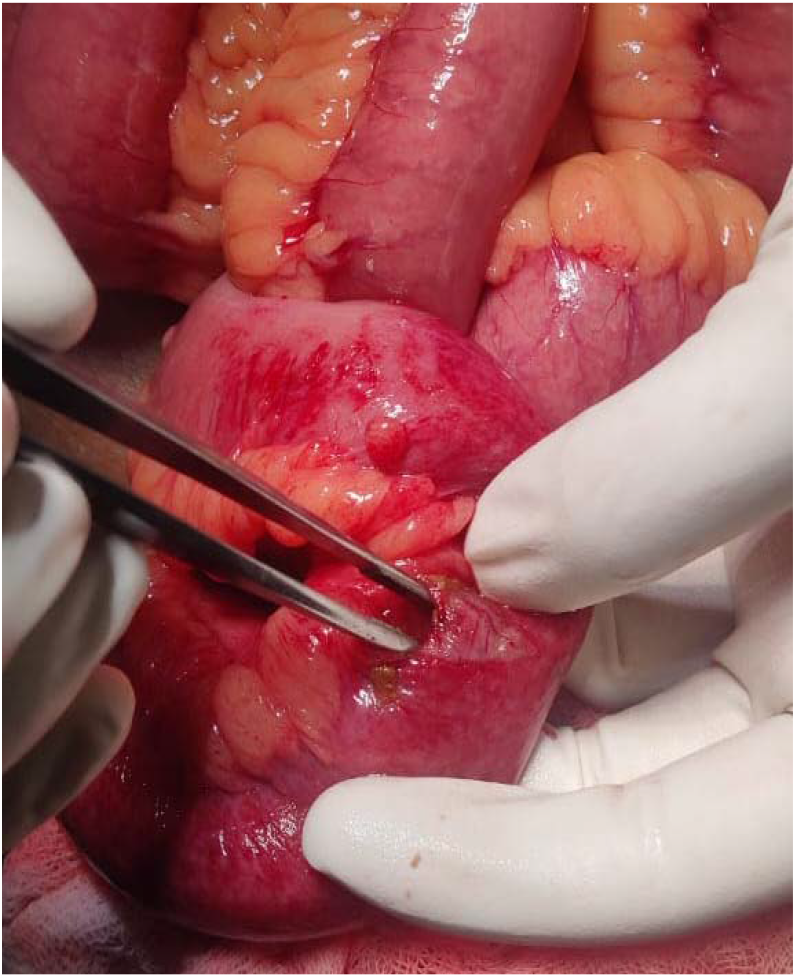
Intraoperative Finding-Ileal perforation.

**Fig 1c:**
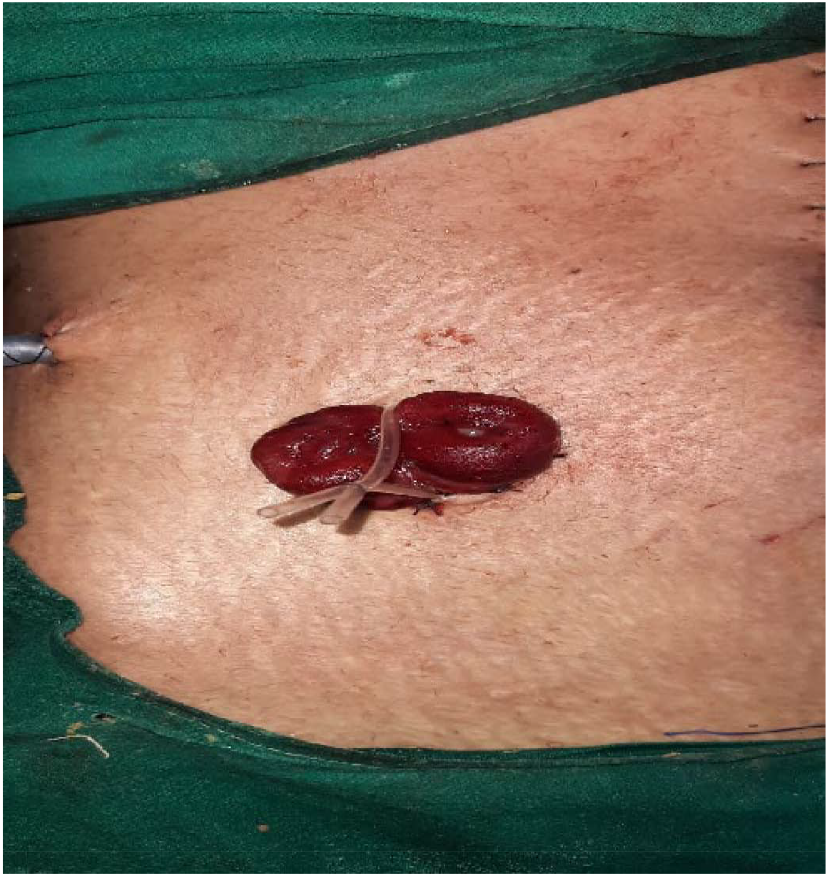
Managed by primary closure of ileal perforation with double barrel ileostomy.

**Fig 2a:**
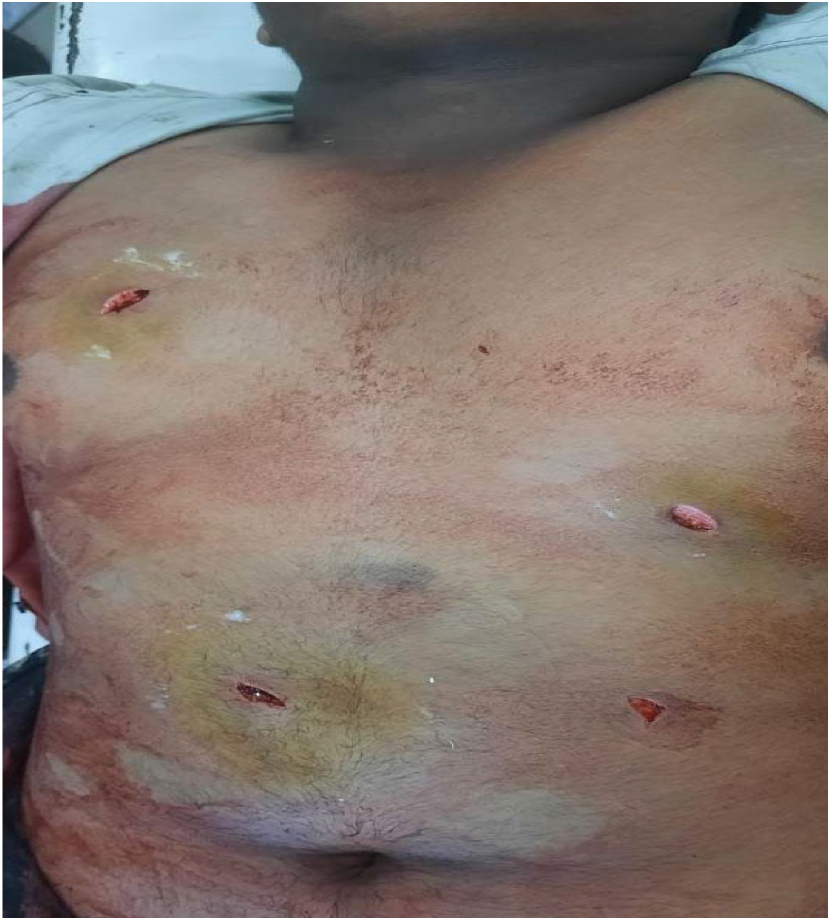
Multiple stab wounds over abdomen and chest.

**Fig 2b:**
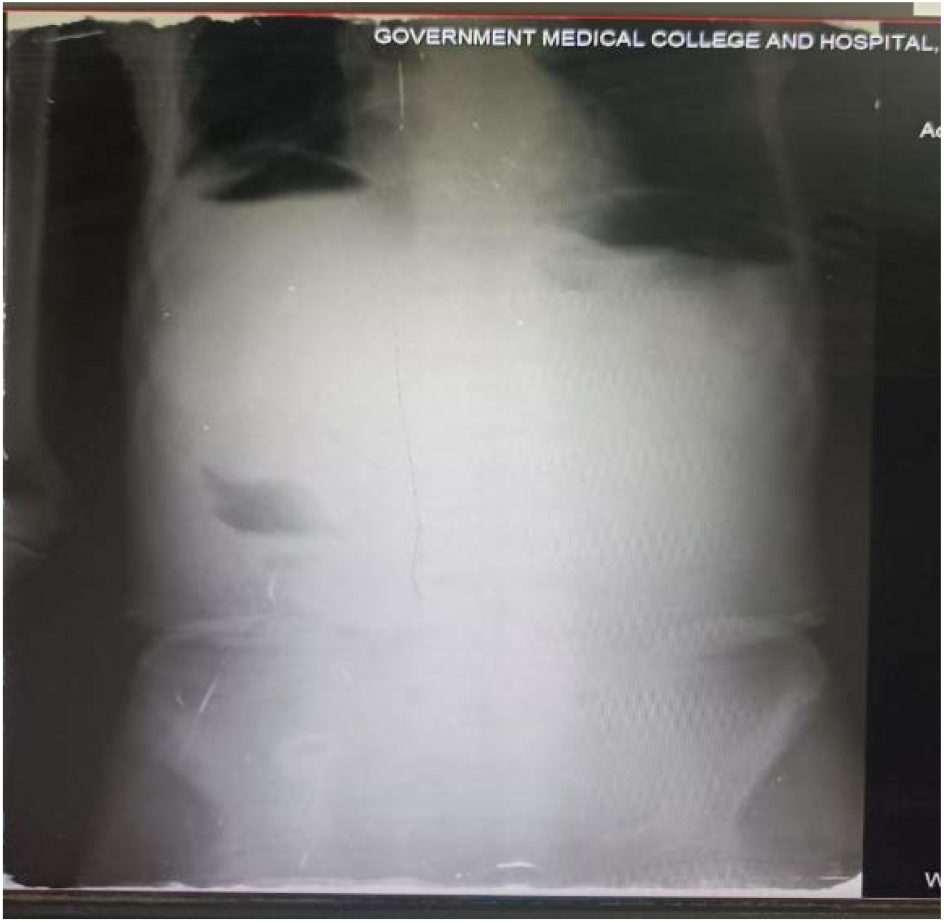
Xray abdomen erect showing air under diaphragm.

**Fig 2c:**
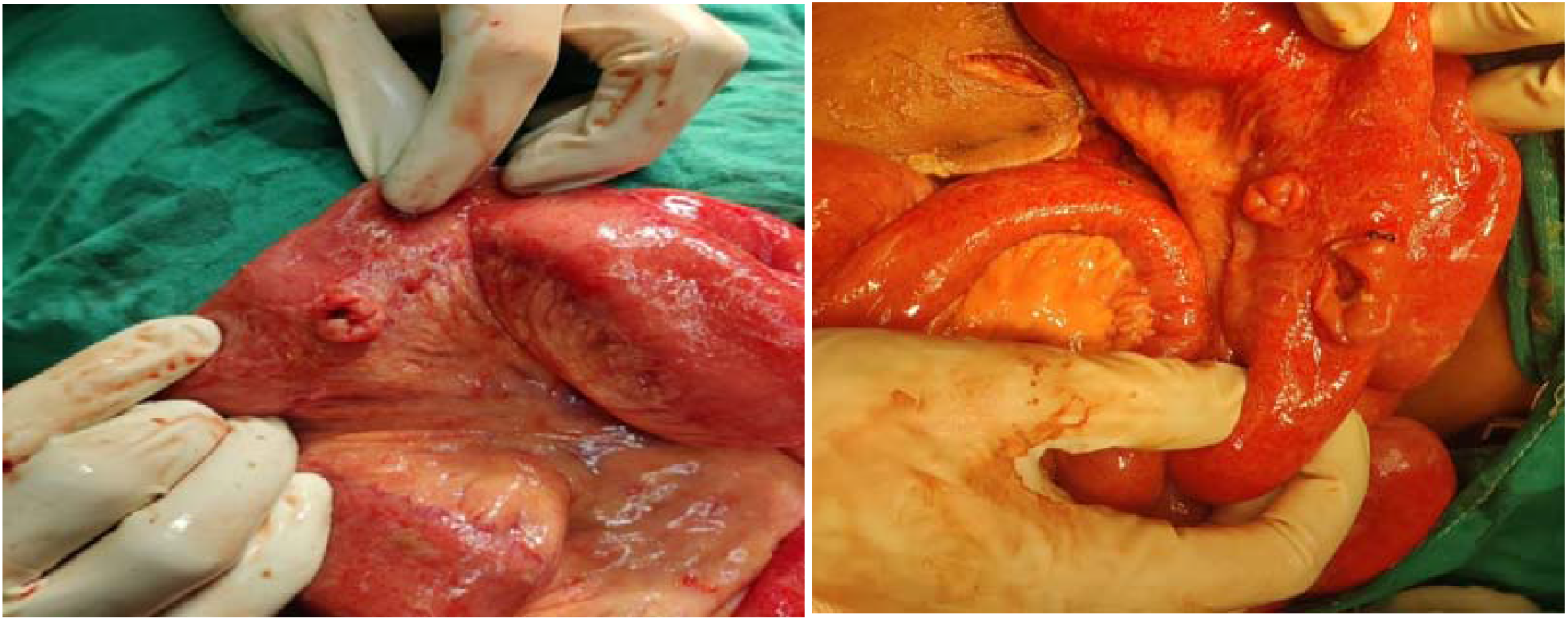
Intraoperative findings: Multiple ileal perforations.

**Fig 2d:**
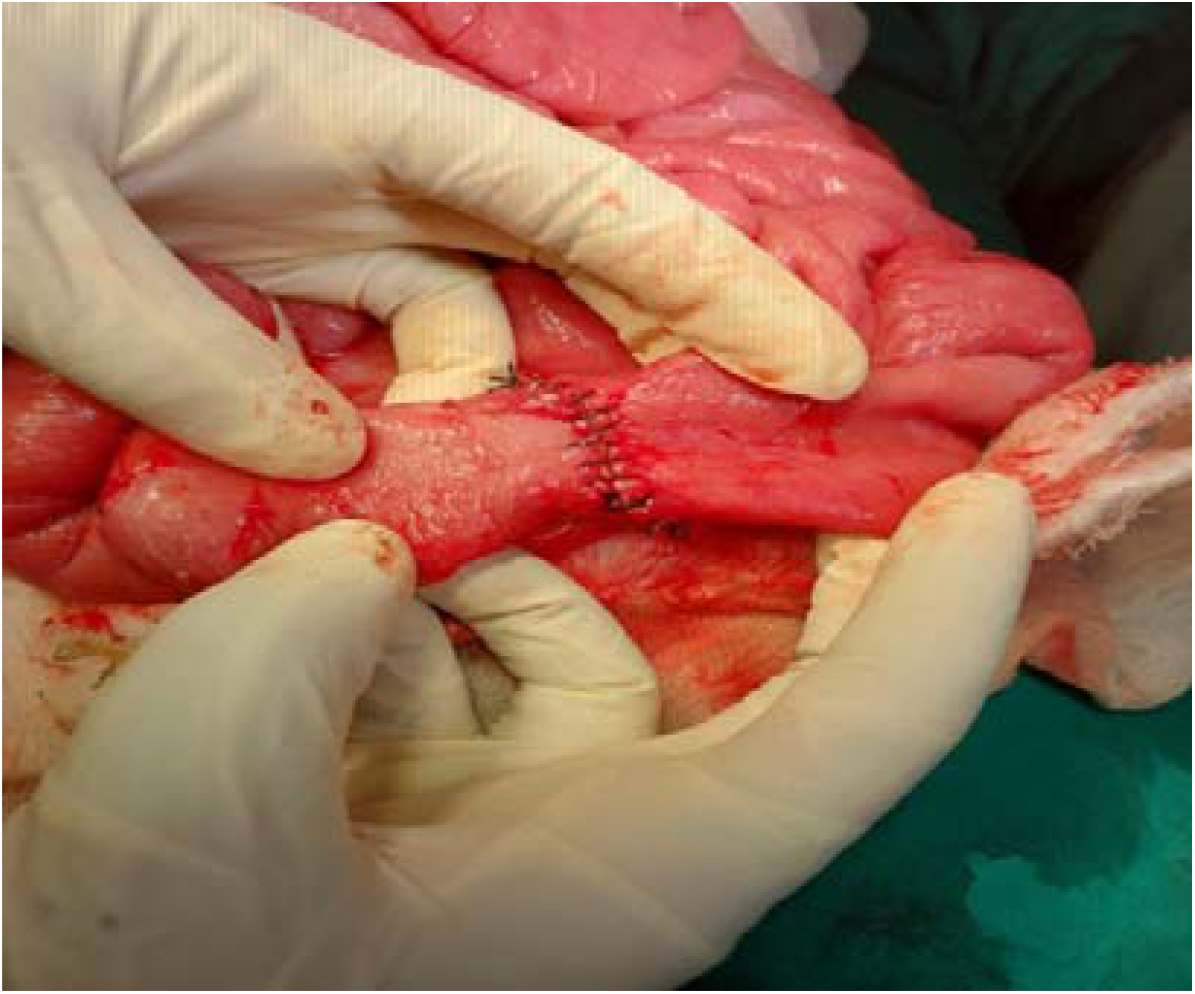
Ileoileal anastomosis.

## Data Availability

All data produced in the present study are available upon reasonable request to the authors

